# Simplified point-of-care full SARS-CoV-2 genome sequencing using nanopore technology

**DOI:** 10.1101/2021.07.08.21260171

**Authors:** Anton Pembaur, Erwan Sallard, Patrick Philipp Weil, Jennifer Ortelt, Parviz Ahmad-Nejad, Jan Postberg

## Abstract

**Background:** The scale of the ongoing SARS-CoV-2 pandemic warrants the urgent establishment of a global decentralized surveillance system to recognize local outbreaks and the emergence of novel variants-of-concern. Among available deep-sequencing technologies, nanopore-sequencing could be an important cornerstone, since it is mobile, scalable and costs-effective. Therefore, streamlined nanopore-sequencing protocols need to be developed and optimized for SARS-CoV-2 variants identification.

**Results:** We adapted and simplified existing workflows using the ‘midnight’ 1,200 bp amplicon split primer sets for PCR, which produce tiled overlapping amplicons covering almost the entire SARS-CoV-2 genome. Subsequently, we applied Oxford Nanopore Rapid Barcoding and the portable MinION Mk1C sequencer combined with the interARTIC bioinformatics pipeline. We tested a simplified and less time-consuming workflow using SARS-CoV-2-positive specimens from clinical routine and identified pre-analytical parameters, which may help to decrease sequencing failures rates. Complete pipeline duration was approx. 7 hrs for one specimen and approx. 11 hrs for 12 multiplexed barcoded specimens.

**Conclusions:** The adapted protocol contains less processing steps and can be completely conducted within one working-day. Diagnostic CT values are principal criteria for specimen selection.

## Background

To face the ongoing SARS-CoV-2 pandemic, a global decentralized warning system is being established to recognize local outbreaks and the emergence of novel variants-of-concern (VOC). A particular focus is given to the identification of VOCs with accelerated transmission rates, increased infectivity or immune escape mutations, since these variants would warrant adaptations in containment and vaccination strategies (Cobey, Larremore et al. 2021, Gupta 2021, Harvey, Carabelli et al. 2021, Winger and Caspari 2021). Moreover, the search for the zoonotic origin of SARS-CoV-2 from comparative analyses of genomic data is an ongoing issue with relevance for the early recognition of future outbreak scenarios (Andersen, Rambaut et al. 2020, V’Kovski, Kzel et al. 2021).

Among the available deep-sequencing technologies, nanopore-sequencing could be an important cornerstone, since it is mobile, scalable and acquisition investments are comparably low. Further, nanopore sequencing devices do not require large-scale IT infrastructure. In the past they were already involved actions of genome surveillance, e.g. the 2015 Ebola outbreak in Liberia, Guinea and sierra Leone (Quick, Loman et al. 2016), and since then there has been a substantial technological progress. However, at least for smaller hospital laboratories with lower throughput, it is desirable to develop protocols as streamlined as possible.

Nanopore sequencing allows the sequencing of either DNA or RNA (Garalde, Snell et al. 2018), and does not require PCR amplification. Furthermore the technique has the potential of producing very long, continuous reads, which theoretically allows to sequence in only one read the 29.903nt long (+)RNA genome of SARS-CoV-2 (Taiaroa, Rawlinson et al. 2020), or its deriving cDNA after reverse transcription (Figure 1).

**Figure 1.**
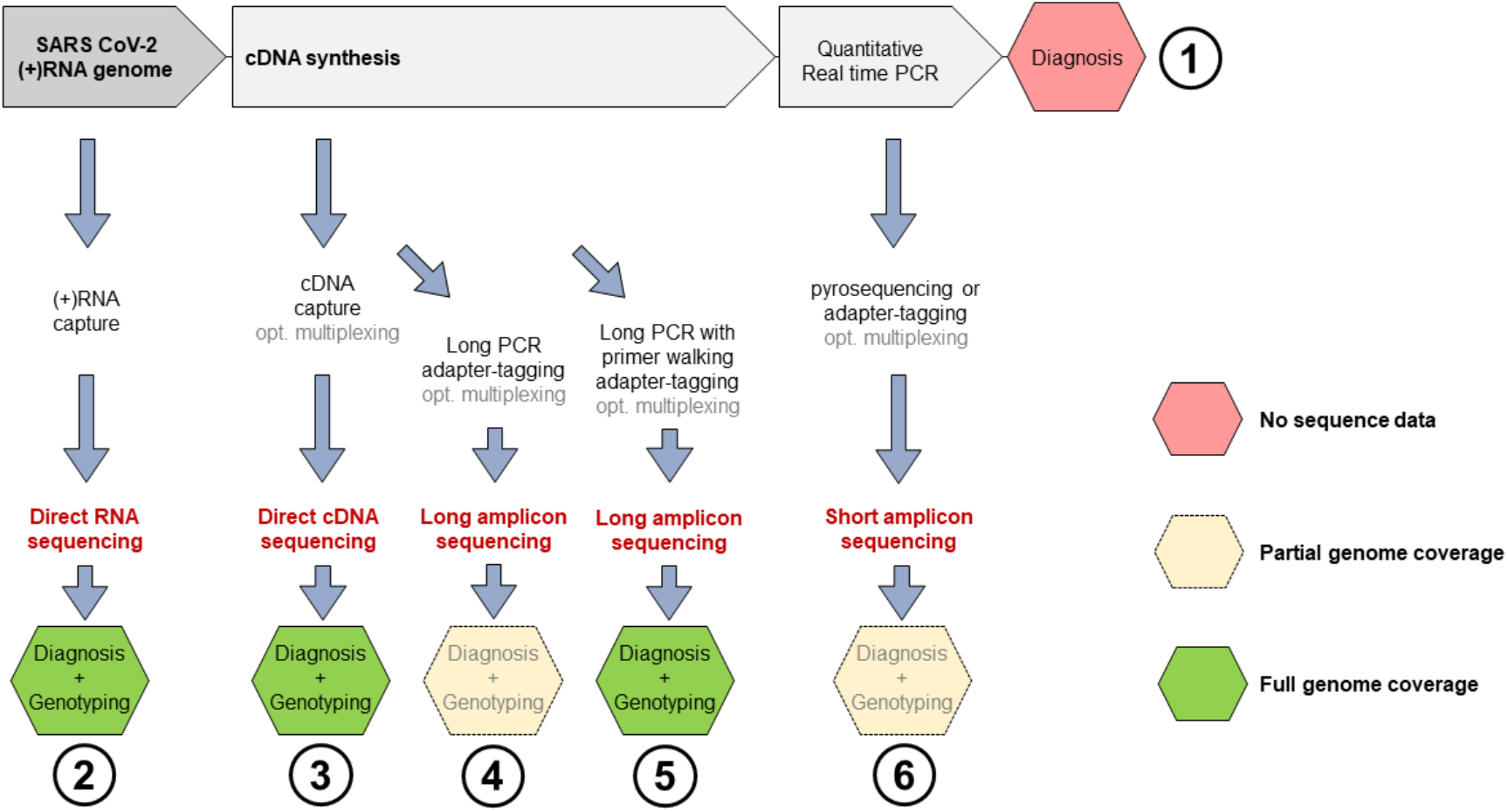
Nanopore sequencing options (2-6) compatible with diagnostic qPCR pipelines (1). The pyrosequencing option in path (6) requires biotin-tagged primers for qPCR (Weil, Hentschel et al. 2021).

With the aims of facilitating implementation in routine diagnostics with lower specimens throughput and of simplifying the workflow, we tested modifications of existing ARTIC protocols for SARS-CoV-2 full length (+)RNA genome sequencing (Freed, Vlkova et al. 2020, Li, Wang et al. 2020, Tyson, James et al. 2020). Furthermore, we tested the simplified and less time-consuming workflow on confirmed SARS-CoV-2-positive specimens from clinical routine and identified pre-analytical parameters, which may help to decrease the rate of sequencing failures.

## Methods

### Nucleic acids isolation

Specimens included nasopharyngeal swabs (Xebios Diagnostics), which underwent routine COVID-19 diagnostic testing. For the collection and use after routine diagnostics procedures we obtained approval of the Witten/Herdecke University Ethics board (No. 160/2020 [For more details, see ‘declarations’ below.]). RNA purification was performed via magnetic beads (Seegene NIMBUS/Tanbead). Alternatively, total RNA was purified from 250 µL liquid specimen using 750 µL QIAzol lysis reagent (Qiagen, Cat. No. 158845) upon manufacturer’s recommendations, or using silica columns (QiaAmp Viral Mini Kit, Qiagen).

### RT-PCR, quality assessments and library preparation

The ‘midnight’ split primer set from the ARTIC protocol was used for SARS-CoV-2 cDNA amplification in 2 multiplex PCR reactions. To avoid overlaps during multiplex PCR, each single-tube PCR reaction generates consecutively tiled, non-overlapping 1,200 bp amplicons. Mixed together after PCR, both resulting complementary amplicon mixtures cover almost the entire SARS-CoV-2 genome (Freed, Vlkova et al. 2020).

Combined reverse transcription and amplification of multiple 1,200 bp amplicons (RT-PCR) was performed in single tube 20 µL reactions using the Luna One-Step RT-qPCR Kit (NEB; E3005). For RT-PCR, 8 µL of purified template RNA were used for each reaction. From 100 µM primer pools, 1 µL was used in each reaction. Reverse transcription was performed at 55°C for 30 minutes, followed by incubation at 95°C for one minute. Then 34 cycles (pool 1) or 30 cycles (pool 2) of denaturation at 95°C for 20 seconds and annealing and extension in one step at 60°C for 210 seconds were performed. A final extension was performed at 65°C. During the implementation phase amplicon sizes and DNA concentrations were routinely checked by agarose gels or by microvolume electrophoresis (Agilent Bioanalyzer, Agilent DNA 12000 kit). Thereafter, amplicons from primer pools 1 and 2 were quantified by fluorimetry (Promega Quantus) and then mixed at equal concentrations. Library preparation was done using the Rapid Barcoding Sequencing Kit (Oxford nanopore; SQK-RBK004) upon manufacturers recommendations.

### Nanopore sequencing

Sequencing was performed on a MinION Mk1C with the options ‘basecalling’ and ‘demultiplexing’ being enabled. As output format, FAST5 and FASTQ files were chosen. Sequencing time was set for 72 hours as default. Sequencing was stopped after reaching at least 10 megabases for each barcode.

### Bioinformatics

Consensus sequences were built from the barcode-sorted, quality-filtered FAST5 and FASTQ files containing sequencing reads, using the interARTIC pipeline. Except otherwise stated, the ‘Nanopolish’ algorithm was routinely used.

Installation and usage of interARTIC pipeline was done following the developers’ instructions: https://psy-fer.github.io/interARTIC/installation/. For faster analysis, all available threads were activated in the advanced settings. The interARTIC pipeline performs read filtering, alignments and returns a consensus FASTA file as well as coverage charts for visualization (Figure 2B-H).

**Figure 2.**
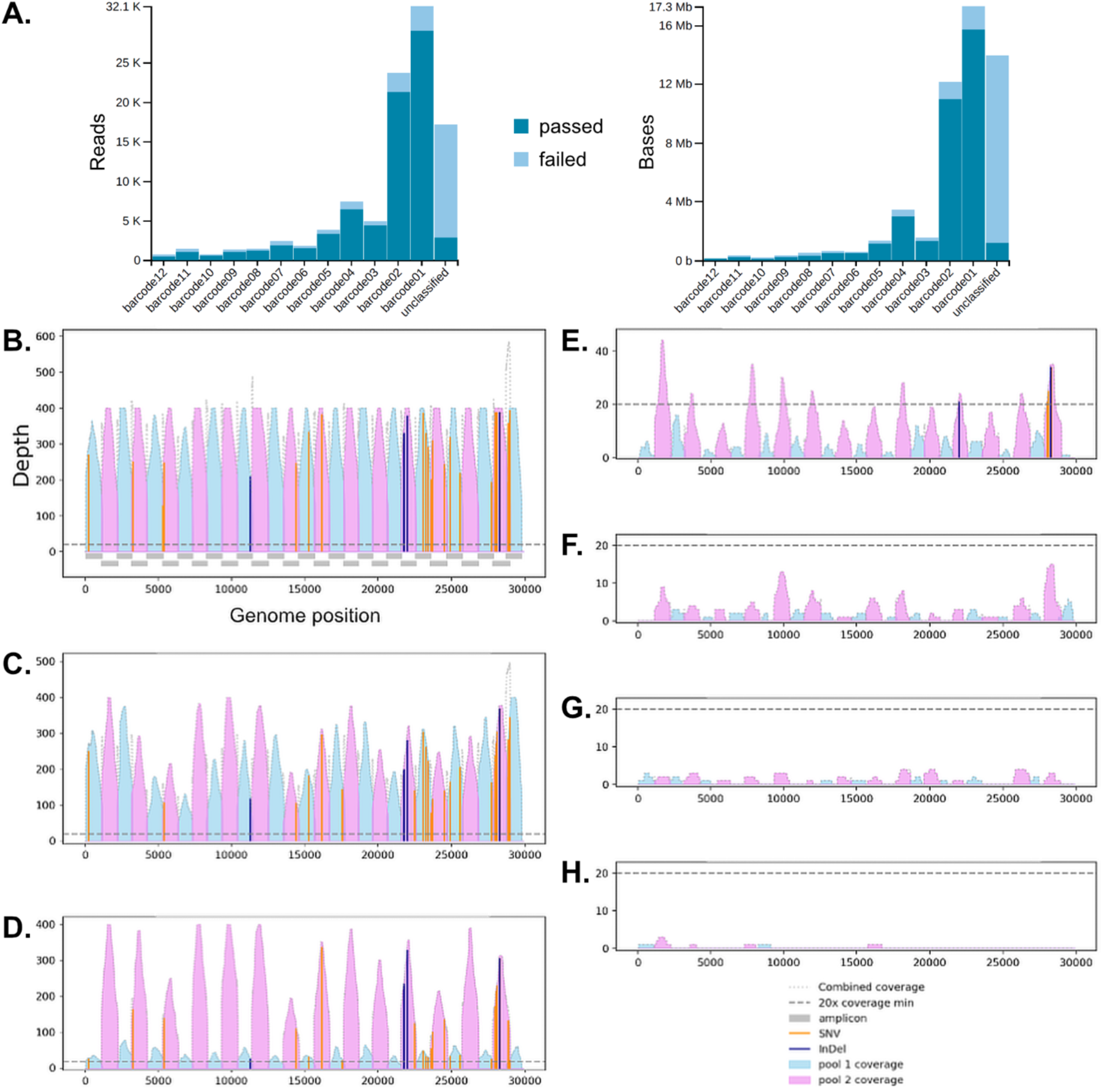
Surveillance using nanopore sequencing (A.), and effects of specimens dilution on the SARS-CoV-2 genome coverage (B.), whereby viral copy numbers were 1.28× 10^6^ (B.); 5.12× 10^8^ (C.); 2.56× 10^8^ (D.); 4× 10^6^ (E.); 2× 10^6^ (F.); 1× 10^6^ (G.); 5× 10^5^ (H.). In (A.) increasing barcode numbers (X-axis) correspond to decreasing viral titers.

Consensus FASTA files were uploaded to the Nextstrain webapp (https://clades.nextstrain.org) to perform phylogenetic analyses (Hadfield, Megill et al. 2018).

### Data availability

FASTQ files and assembled FASTA formatted consensus sequences are available (https://t1p.de/minion-seqdata)

## Results and Discussion

We defined milestones to simplify the protocol and decrease hands-on sample time and working step numbers. 1. We tested whether specimens can be directly taken from residual diagnostic specimens extracted from 96-deepwell-plates using magnetic beads (Seegene NIMBUS/Tanbead). For comparison we applied RNA purification protocols using silica columns (QiaAmp Viral Mini Kit, Qiagen) or guanidinium isothiocyanate (GITC) for RNA extraction (QIAzol lysis reagent, Qiagen). 2. We tested whether reverse transcription can be successfully primed by SARS-CoV-2-specific primers, which are subsequently used for multiplex 1,200 bp amplicon amplification. Purified RNA from the different extraction protocols of milestone 1 was used for cDNA synthesis and successive multiplex PCR with the 2 ‘midnight’ split 1,200 bp amplicon primer pools in single tube reactions 3. We aimed at improving the efficiency of the nanopore sequencing workflow by using the onboard Guppy basecalling capability of the Oxford Nanopore MinION Mk1C device. Moreover, implementation of the interARTIC interface should help to avoid command line-based bioinformatics analyses as much as possible in order to provide a user-friendly and efficient analysis pipeline.

### Protocol implementation considering differences in viral loads and influences of RNA extraction protocols

Sequencing was performed from serially diluted specimens of purified RNA from patients’ samples, which exhibited low cycle threshold (CT) value (CT=16-18) after routine diagnostics RT-qPCR (N gene, RdRP gene; Seegene). Using the quantitative reference sample Ch07470 for calibration we determined that a CT=25 (respectively 16 and 18) corresponded to a SARS-CoV-2 copy number of 1.0× 10^6^ (respectively 5.12× 10^8^ and 1.28× 10^8^). Routinely used dilution factors were 2^0^-2^-10^ in order to cover several CT value magnitudes and to simulate different amounts of viral loads. In terms of RNA yield and 1,200 bp amplicon PCR performance the magnetic beads-based RNA purification protocol outperformed slightly the GITC method as well as the column-based protocol, but we did not observe differences in read and coverage quality between these different isolation methods (Figure S1). Since direct sampling from 96-deepwell plates allowed us to directly exploit residual specimens, which remained after routine RT-qPCR diagnostics, we decided to focus on magnetic beads-based RNA purification during further protocol development. Moreover, it shortened and simplified the workflow. After semiquantitative or quantitative RT-PCR using multiplex primer pools 1 and 2 in separate single tube reactions for combined reverse transcription of the SARS-CoV-2 (+)RNA and amplification of 1,200 bp amplicons, band intensities exhibited strong dependence on viral loads. Moreover, after 32 PCR cycles band intensities using primer pool 1 were weaker when compared with primer pool 2 (Figure S2). We determined that 34 PCR cycles for primer pool 1 and 30 cycles for primer pool 2 were a good compromise. Therefore, reverse transcription can be successfully primed by the ‘midnight’ primers.

### Surveillance of multiplex nanopore sequencing and multiple reuses of flow cells

Using the Rapid Barcoding Kit (SQK-RBK004, Oxford Nanopore) enabled us to barcode the PCR products without purification steps and sequence them immediatley. As a standard, we used 12 barcoded libraries for multiplex sequencing on R9 flow cells. In contrast to the Oxford Nanopore MinION Mk1B, the MinION Mk1C device features onboard guppy basecalling. In combination with the Rapid Barcoding Kit, we exploited this opportunity for real-time surveillance of basecalling and demultiplexing for each of the 12 multiplexed samples per run. This enabled us to recognize the exact time point at which a reading depth of approx. 10 Mbp per barcode was achieved. The time to reach this threshold depended heavily on the viral load (simulated by serially diluted samples). Decreased viral loads let to a considerable decrease of passed reads (Figure 2A). A manual stop of sequencing followed by flow cell washing (Flow Cell Wash Kit, EXP-WSH004, Oxford Nanopore) allowed us to reuse a single flow cell for a series of 3 sequencing runs, each using 12 multiplexed barcoded libraries. This specific scenario resulted in costs of approx. 40 USD per sample. Theoretically, the possible number of reuses depends largely on the duration of the sequencing, which itself depends mainly on the number of used barcodes. Further, if a MinION Mk1B is considered for this purpose, a similar surveillance functionality could be achieved using RAMPART (https://github.com/artic-network/rampart) on a dedicated LINUX environment.

### Assembly of full SARS-CoV-2 genomes and pathogen genome data analyses

For mapping and full length SARS-CoV-2 genome assembly, we used the FASTQ files resulting from Guppy basecalling in order to generate FASTA formatted consensus sequence files. We used the ARTIC pipeline through a graphical user interface (interARTIC; https://github.com/Psy-Fer/interARTIC). Once installed, this is an easy to use and relatively fast pipeline with only five minutes hands-on time, which enables the use of the ‘Nanopolish’ or ‘Medaka’ algorithms for simultaneous analyses of multiplexed barcoded samples. As an example, Figure 2B shows the complete and deep coverage of the complete SARS-CoV-2 genome after the combination of pools 1 (light blue) and 2 (pink). This demonstrates that all contained 1,200 bp amplicon were specifically and efficiently amplified during the combined RT-PCR reaction (Figure 2B).

To compare the interARTIC and Geneious Prime pipelines, we used exactly the same FASTQ file from the same sample shown in Figure 2B (https://go.geneious.com/video/how-to-assemble-coronavirus-genomes). Phylogenetic analyses with Nextstrain using the FASTA consensus files obtained from the interARTIC or the Geneious Prime pipelines resulted in considerably different phylogenetic distances in clade 20I, showing that the bioinformatics pipeline influences the result (Figure S3). The interARTIC pipeline proved superior in terms of coverage and sequencing depth. Notably, within the interARTIC pipeline both options, the ‘Nanopolish’ and ‘Medaka’ algorithms performed equally well with respect to consensus sequence quality, but ‘Medaka’ was considerably faster.

We observed that the consensus sequences returned by the ‘Nanopolish’ and ‘Medaka’ algorithms contain numerous unsolved regions (for which only ‘N’s are indicated). Interestingly, the regions unsolved by one algorithm were generally solved by the other, so we assumed that the two consensus sequences could be combined to produce a ‘super-consensus’ with improved variant prediction value. We developed a Python code that merges the ‘Nanopolish’ and ‘Medaka’ consensus sequences and generates the corresponding variant calling file. As expected, the ‘super-consensus’ contained less unsolved regions when compared with the ‘Nanopolish’ and ‘Medaka’ consensus sequences alone (Figure 4B). In addition, it retained the high quality mutations, which were identified by both algorithms, while removing most false-positives probably caused by sequencing and alignment errors (Figure 4A). Consequently, our consensus-merging code improves the quality of variant calling and highlights the complementarity of ‘Nanopolish’ and ‘Medaka’ for nanopore-sequencing of SARS-CoV-2 and others. Nevertheless, this quality increase comes at the cost of information loss such as the number of reads per variant or other metadata which were initially generated by ‘Nanopolish’ and ‘Medaka’ and are not transferred to the ‘super-consensus’.

Serial input RNA dilutions or, respectively, viral load influenced the depth of sequencing (Figure 2C-H). For the output of high-quality consensus sequences in the FASTA file format a coverage threshold of 20 was used as default. We generally observed that this could be reached when a SARS-CoV-2 titer of 4× 10^6^ was given. Viral copy numbers lower than 4× 10^6^ were associated with incompletely assembled SARS-CoV-2 genomes. Thus, we provide here a convincing line of evidence that the copy number-normalized CT values of diagnostic RT-qPCR can be used as the criterion of sequencing success.

Single or batch high-quality consensus FASTA formatted sequences were used for phylogenetic tree visualization and variant calling using the Nextstrain webapp (https://clades.nextstrain.org) (Hadfield, Megill et al. 2018). In our hands, the obtained sequences could faithfully be assigned to specific clades in the reference tree (Figure 3). Again, an influence of viral load was observed. However, despite incomplete coverage in those cases enough informative sequence data could be obtained for phylogenetic analyses from several low copy number samples. As a result of serial sample dilutions, we observed deviating phylogenetic distances within the clade, wherein specimens classification occurred (Figure 3A), which eventually could lead to incorrect clade association. The use of specimens from diagnostic routine with viral copy numbers higher than approx. 4× 10^6^ apparently led to their faithful association with different clades, which were clades 20I and 19A in the shown example (Figure 3B). The ‘Nanopolish’ and ‘Medaka’ consensus as well as the merged super-consensus could be reliably associated to the corresponding clade (20I in the shown example). The ‘Medaka’ consensus mapped at a greater distance than the ‘Nanopolish’ consensus, probably because ‘Medaka’ algorithm does not correct frameshifts, while the super-consensus had an intermediary distance between the two other consensus (Figure 4C).

**Figure 3.**
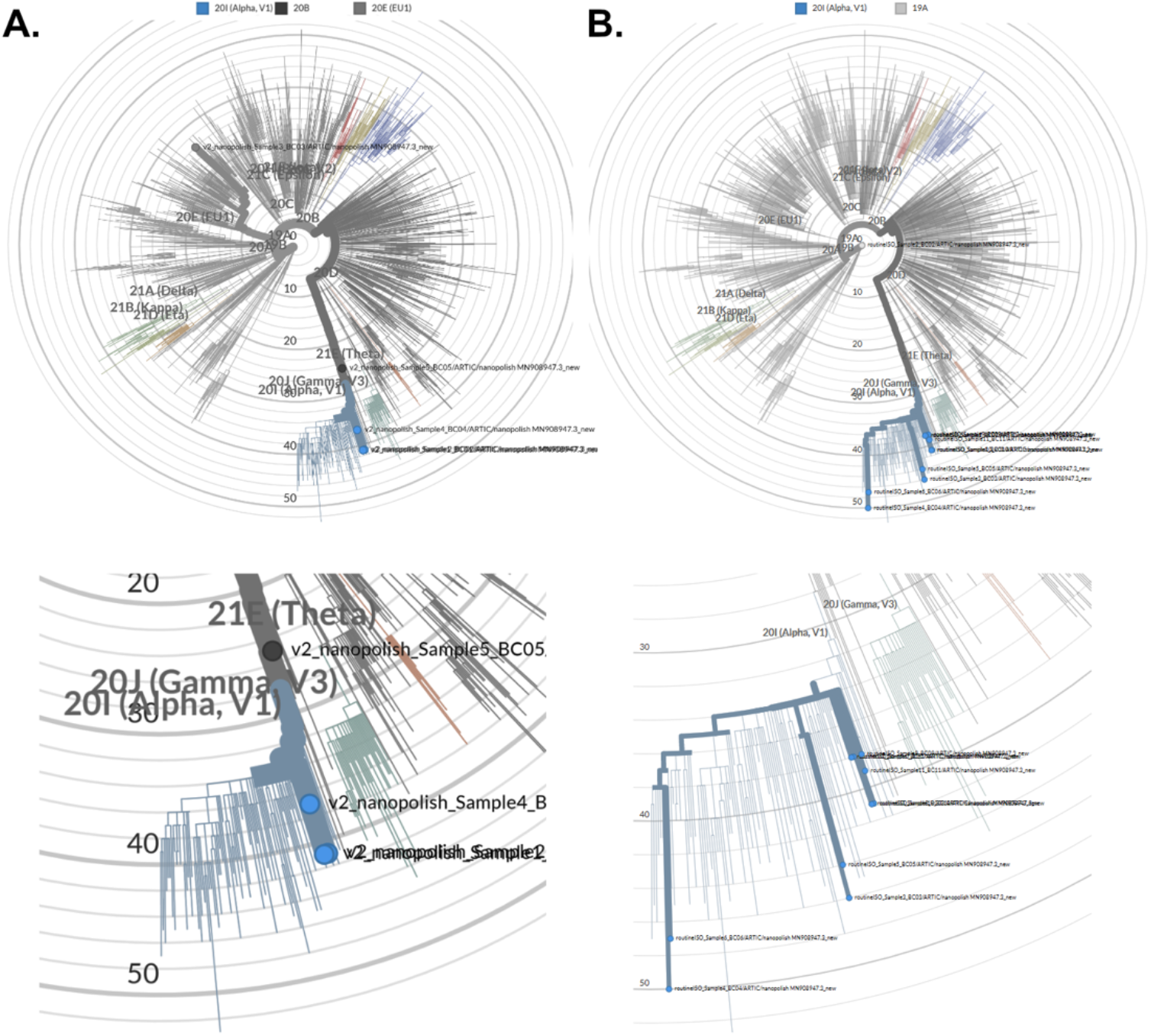
Comparison on the evolutionary distance calculation, when data from the same specimen was processed by 2 different bioinformatics pipelines. Phylogenetic tree visualization was done using the Nextstrain open-source platform for pathogen genome data analyses (Hadfield, Megill et al. 2018). We used serial dilutions of identical specimens (A.), or a selection of samples from different individuals from clinical routine (B.) for phylogenetic analyses. Below the trees a magnification of clade 20I is shown, wherein most specimens grouped (A.,B.).

**Figure 4.**
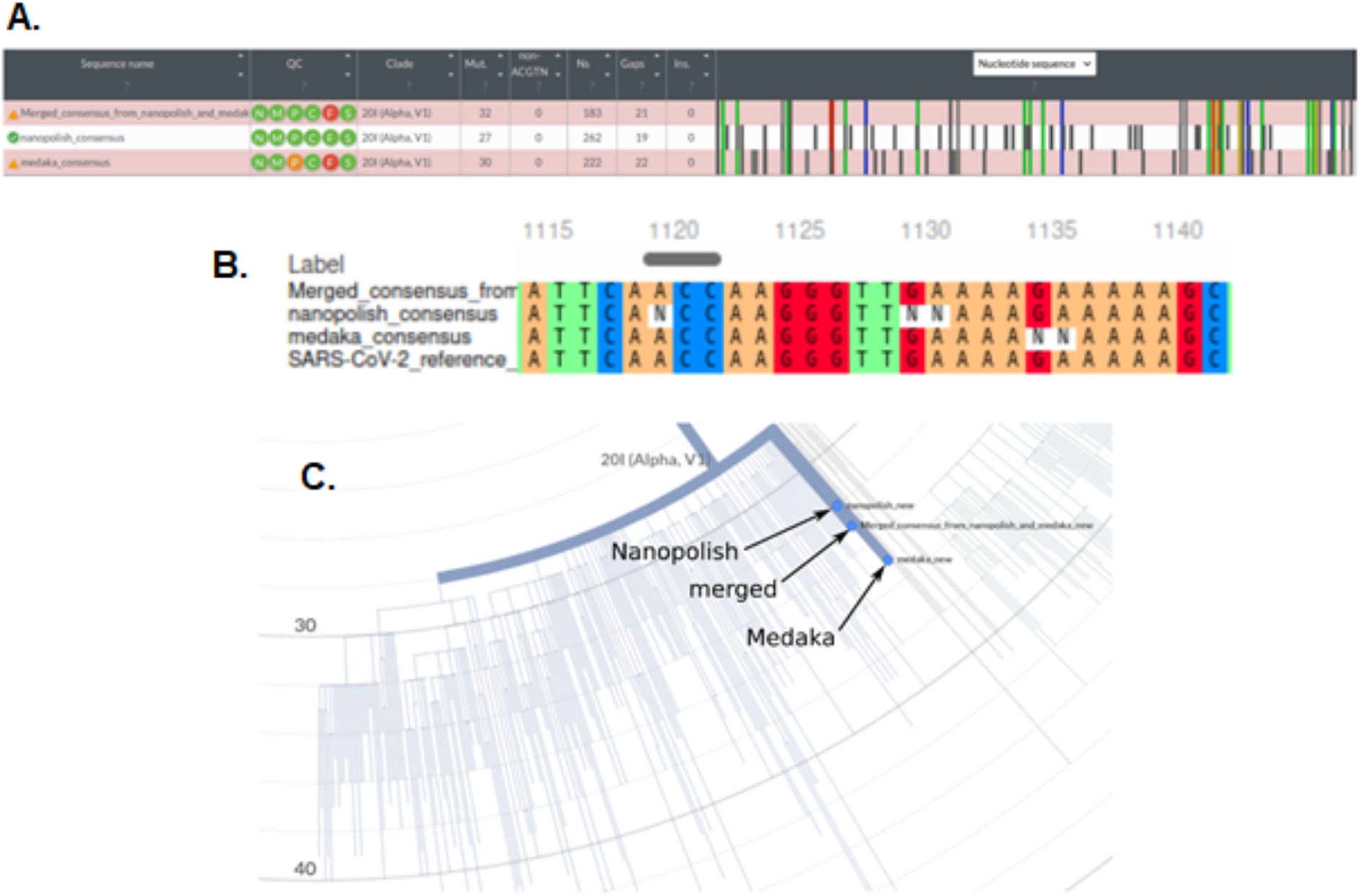
Comparison of the consensus sequences returned by ‘Nanopolish’ and ‘Medaka’ algorithm and the merged consensus for a nanopore sequencing run corresponding to a viral load of 1.28×10**6. **(A.)** Alignment of the ‘Nanopolish’ and ‘Medaka’ consensus as well as the merged consensus on the SARS-CoV-2 reference genome by the Nextstrain program. The merged consensus conserved all of the high quality mutations that mapped to the known variant 20I (shown in colour or light grey), while most non-matching mutations (in dark grey, likely sequencing errors) of the ‘Nanopolish’ or ‘Medaka’ consensus were lost. **(B.)** Comparison of variant solving by ‘Nanopolish’, ‘Medaka’ and our code. The multiple alignment was performed by MAFFT online tool with the three consensus sequences and the SARS-CoV-2 reference genome (MN908947.3). Contrary to the ‘Nanopolish’ and ‘Medaka’ consensus, the merged consensus solved the entire region and led to accurate variant calling. **(C.)** Clade mapping and phylogenetic distances calculated by Nextstrain for the three consensus sequences.

Taken together, the main achievements of an optimized workflow are: 1. Purified RNA from SARS-CoV-2-positive patients can be directly taken from residual diagnostic specimens in 96-deepwell-plates; 2. cDNA synthesis and successive multiplex PCR with 2 split primer pools can be performed in single tube reactions. Since cDNA synthesis is primed by SARS-CoV-2-specific primers for 1,200 bp amplicon amplification, there is no need for use of unspecific hexanucleotide priming. 3. Onboard Guppy basecalling with the Oxford Nanopore MinION Mk1C device and implementation of the interARTIC led to a further reduction of working steps and hands-on time (Figure 5). Implementation in smaller hospital laboratories with lower specimens’ throughput can be easily done at moderate costs.

**Figure 5.**
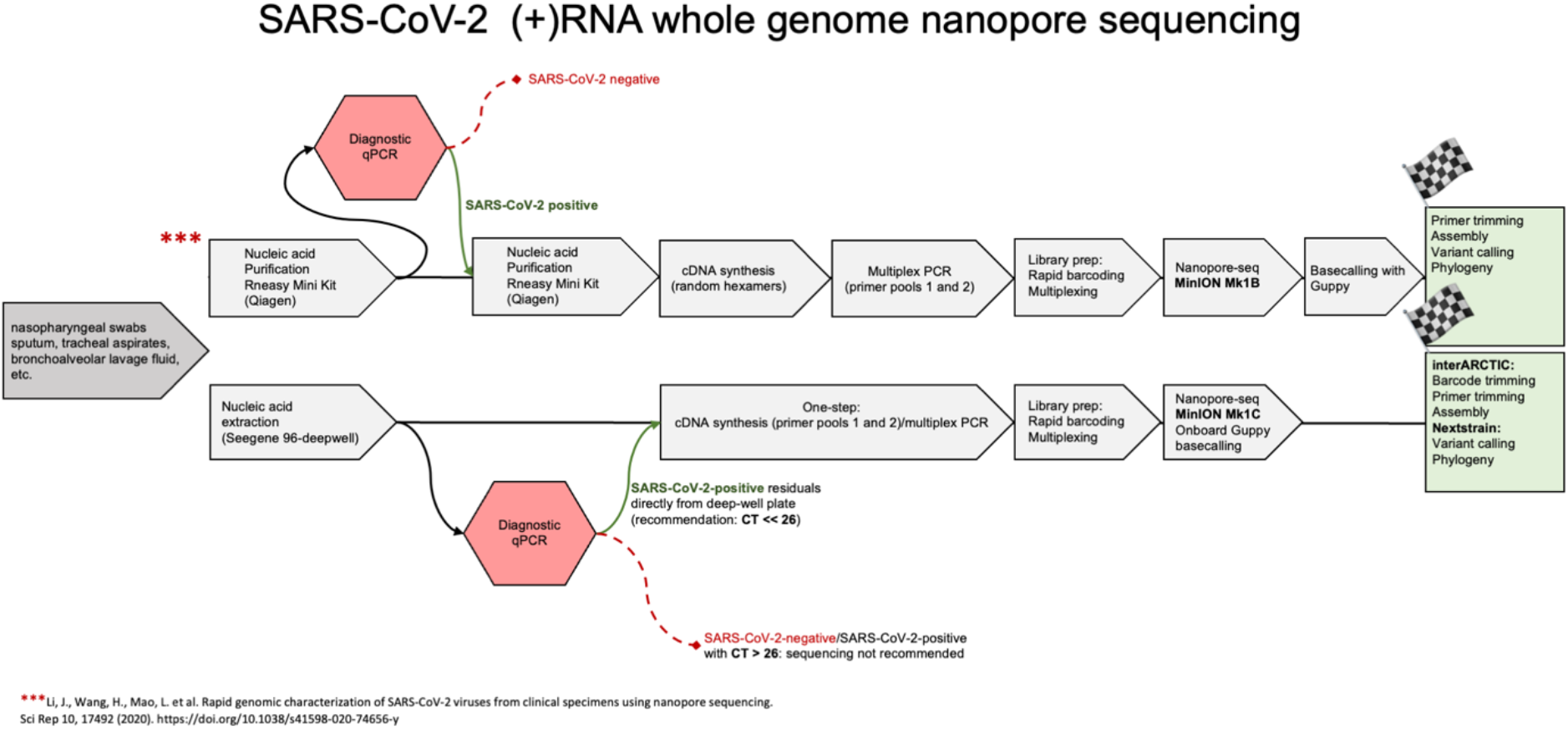
Comparison of a rapid SARS-CoV-2 whole (+)RNA genome nanopore sequencing pipeline (Freed, Vlkova et al. 2020) with the novel simplified workflow, whose main achievements are: 1. Purified RNA from SARS-CoV-2-positive patients can be directly taken from residual diagnostic specimens in 96-deepwell-plates; 2. cDNA synthesis and successive multiplex PCR with 2 primer pools can be performed in single tube reactions. Since cDNA synthesis is primed by SARS-CoV-2-specific primers for 1,200 bp amplicon amplification, there is no need for use of unspecific hexanucleotide priming. 3. Onboard Guppy basecalling with the Oxford Nanopore MinION Mk1C device and implementation of the interARTIC led to a further reduction of working steps and hands-on time.

We provide a detailed protocol for all steps here, which includes the Python code (Supplementary Information 1) and corresponding command line: <u>dx.doi.org/10.17504/protocols.io.bwhppb5n</u>

## Conclusions

The adapted protocol contains less processing steps. Diagnostic CT values are the principal criteria for specimen selection. After diagnostic qRT-PCR, multiplex library preparation, quality controls, nanopore sequencing and the bioinformatic pipeline can be completely conducted within one working-day.

## Supporting information

Figures S1-3 and Supplementary Information 1 (Python code)

## Data Availability

FASTQ files and assembled FASTA formatted consensus sequences are available (https://t1p.de/minion-seqdata).
We provide a detailed protocol here, which includes the Python code and corresponding command line: dx.doi.org/10.17504/protocols.io.bvstn6en

## List of abbreviations

(+)RNA: (sense polarity [single-stranded] ribonucleic acid)
RdRP: (RNA-dependent RNA polymerase)
RT-qPCR: (reverse transcription-quantitative polymerase chain reaction
SARS-CoV-2: (severe acute respiratory syndrome coronavirus 2)

## Declarations

### Ethics approval and consent to participate

For the collection and use of specimens from clinical routine at Helios University Hospital Wuppertal (North Rhine-Westphalia, Western Germany) we obtained approval of the Witten/Herdecke University Ethics board (No. 160/2020). The positive ethics vote included the use of routinely sampled and SARS-CoV-2-confirmed specimens, which underwent COVID-19 RT-qPCR diagnostics procedures even for cases, where informed written consent could not be asked for. When possible, we obtained informed written consent from hospitalized patients or legal guardians. All work has been conducted according to the principles expressed in the Declaration of Helsinki.

## Authors’ contributions

AP prepared libraries for nanopore-seq and conducted the sequencing. PAN and JO collected specimens and conducted diagnostic SARS-CoV-2 qPCR. AP and PPW performed nucleic acids quality controls. AP, ES and JP contributed to the bioinformatics analyses. ES developed the Python code. PAN and JP designed the sudy. AP, ES, PPW and JP wrote the paper.

## References

Andersen, K. G., A. Rambaut, W. I. Lipkin, E. C. Holmes and R. F. Garry (2020). “The proximal origin of SARS-CoV-2.” Nat Med 26(4): 450–452.

Cobey, S., D. B. Larremore, Y. H. Grad and M. Lipsitch (2021). “Concerns about SARS-CoV-2 evolution should not hold back efforts to expand vaccination.” Nat Rev Immunol 21(5): 330–335.

Freed, N. E., M. Vlkova, M. B. Faisal and O. K. Silander (2020). “Rapid and inexpensive whole-genome sequencing of SARS-CoV-2 using 1200 bp tiled amplicons and Oxford Nanopore Rapid Barcoding.” Biol Methods Protoc 5(1): bpaa014.

Garalde, D. R., E. A. Snell, D. Jachimowicz, B. Sipos, J. H. Lloyd, M. Bruce, N. Pantic, T. Admassu, P. James, A. Warland, M. Jordan, J. Ciccone, S. Serra, J. Keenan, S. Martin, L. McNeill, E. J. Wallace, L. Jayasinghe, C. Wright, J. Blasco, S. Young, D. Brocklebank, S. Juul, J. Clarke, A. J. Heron and D. J. Turner (2018). “Highly parallel direct RNA sequencing on an array of nanopores.” Nat Methods 15(3): 201–206.

Gupta, R. K. (2021). “Will SARS-CoV-2 variants of concern affect the promise of vaccines?” Nat Rev Immunol 21(6): 340–341.

Hadfield, J., C. Megill, S. M. Bell, J. Huddleston, B. Potter, C. Callender, P. Sagulenko, T. Bedford and R. A. Neher (2018). “Nextstrain: real-time tracking of pathogen evolution.” Bioinformatics 34(23): 4121–4123.

Harvey, W. T., A. M. Carabelli, B. Jackson, R. K. Gupta, E. C. Thomson, E. M. Harrison, C. Ludden, R. Reeve, A. Rambaut, C.-G. U. Consortium, S. J. Peacock and D. L. Robertson (2021). “SARS-CoV-2 variants, spike mutations and immune escape.” Nat Rev Microbiol 19(7): 409–424.

Li, J., H. Wang, L. Mao, H. Yu, X. Yu, Z. Sun, X. Qian, S. Cheng, S. Chen, J. Chen, J. Pan, J. Shi and X. Wang (2020). “Rapid genomic characterization of SARS-CoV-2 viruses from clinical specimens using nanopore sequencing.” Sci Rep 10(1): 17492.

Quick, J., N. J. Loman, S. Duraffour, J. T. Simpson, E. Severi, L. Cowley, J. A. Bore, R. Koundouno, G. Dudas, A. Mikhail, N. Ouedraogo, B. Afrough, A. Bah, J. H. Baum, B. Becker-Ziaja, J. P. Boettcher, M. Cabeza-Cabrerizo, A. Camino-Sanchez, L. L. Carter, J. Doerrbecker, T. Enkirch, I. G. G. Dorival, N. Hetzelt, J. Hinzmann, T. Holm, L. E. Kafetzopoulou, M. Koropogui, A. Kosgey, E. Kuisma, C. H. Logue, A. Mazzarelli, S. Meisel, M. Mertens, J. Michel, D. Ngabo, K. Nitzsche, E. Pallash, L. V. Patrono, J. Portmann, J. G. Repits, N. Y. Rickett, A. Sachse, K. Singethan, I. Vitoriano, R. L. Yemanaberhan, E. G. Zekeng, R. Trina, A. Bello, A. A. Sall, O. Faye, O. Faye, N. Magassouba, C. V. Williams, V. Amburgey, L. Winona, E. Davis, J. Gerlach, F. Washington, V. Monteil, M. Jourdain, M. Bererd, A. Camara, H. Somlare, A. Camara, M. Gerard, G. Bado, B. Baillet, D. Delaune, K. Y. Nebie, A. Diarra, Y. Savane, R. B. Pallawo, G. J. Gutierrez, N. Milhano, I. Roger, C. J. Williams, F. Yattara, K. Lewandowski, J. Taylor, P. Rachwal, D. Turner, G. Pollakis, J. A. Hiscox, D. A. Matthews, M. K. O’Shea, A. M. Johnston, D. Wilson, E. Hutley, E. Smit, A. Di Caro, R. Woelfel, K. Stoecker, E. Fleischmann, M. Gabriel, S. A. Weller, L. Koivogui, B. Diallo, S. Keita, A. Rambaut, P. Formenty, S. Gunther and M. W. Carroll (2016). “Real-time, portable genome sequencing for Ebola surveillance.” Nature 530(7589): 228–232.

Taiaroa, G., D. Rawlinson, L. Featherstone, M. Pitt, L. Caly, J. Druce, D. Purcell, L. Harty, T. Tran, J. Roberts, M. Catton, D. Williamson, L. Coin and S. Duchene (2020). “Direct RNA sequencing and early evolution of SARS-CoV-2.” bioRxiv: 2020.2003.2005.976167.

Tyson, J. R., P. James, D. Stoddart, N. Sparks, A. Wickenhagen, G. Hall, J. H. Choi, H. Lapointe, K. Kamelian, A. D. Smith, N. Prystajecky, I. Goodfellow, S. J. Wilson, R. Harrigan, T. P. Snutch, N. J. Loman and J. Quick (2020). “Improvements to the ARTIC multiplex PCR method for SARS-CoV-2 genome sequencing using nanopore.” bioRxiv: 2020.2009.2004.283077.

V’Kovski, P., A. Kratzel, S. Steiner, H. Stalder and V. Thiel (2021). “Coronavirus biology and replication: implications for SARS-CoV-2.” Nat Rev Microbiol 19(3): 155–170.

Weil, P. P., J. Hentschel, F. Schult, A. Pembaur, B. Ghebremedhin, O. Mboma, A. Heusch, A. C. Reuter, D. Muller, S. Wirth, M. Aydin, A. C. W. Jenke and J. Postberg (2021). “Combined RT-qPCR and pyrosequencing of a Spike glycoprotein polybasic cleavage motif can uncover pediatric SARS-CoV-2 infections associated with heterogeneous presentation.” Mol Cell Pediatr 8(1): 4.

Winger, A. and T. Caspari (2021). “The Spike of Concern-The Novel Variants of SARS-CoV-2.” Viruses 13(6).

