## Supplementary material for "Simplified point-of-care full SARS-CoV-2 genome sequencing using nanopore technology": Figures S1-3 and Supplementary Information 1 (Python code)

| | |
|---|---|
| Anton Pembaur: | |
| Erwan Sallard: | |
| Patrick Philipp Weil: | |
| Jennifer Ortelt: | |
| Parviz Ahmad-Nejad: | |
| Jan Postberg: | |

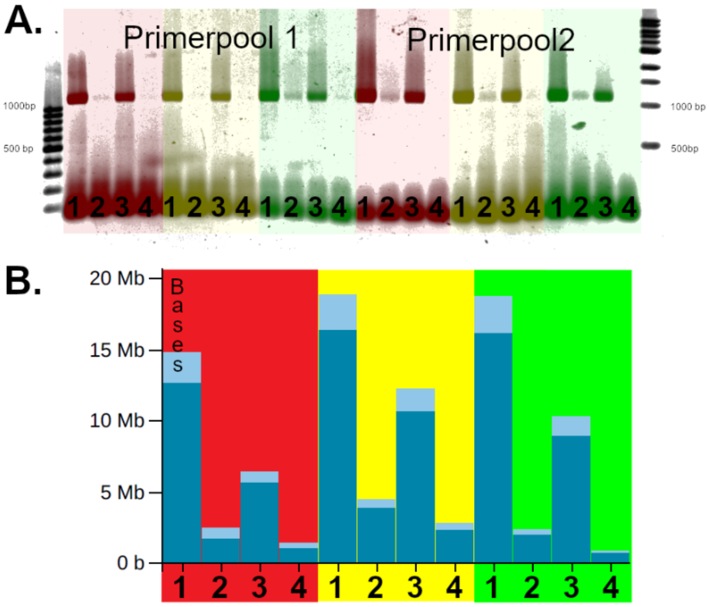


**Figure S1.** Comparison of different isolation methods with respect to PCR performance (**A**.) and nanopore sequencing outcome (**B**.): Silica columns (red shaded), magnetic beads (yellow shaded) ore GITC (green shaded) were used for RNA extraction. Four samples with different Ct-Values were used (1: Ct 19, 2: Ct 25, 3: Ct 19, 4: Ct 24). **A.:** Results of agarose gel electrophoresis after 1,200 bp amplicon generating multiplex PCR. The image shows the uncropped agarose gel. After image acquisition the colors were inverted for better visualization. Thereafter color shades were overlaid to facilitate sample group identification.

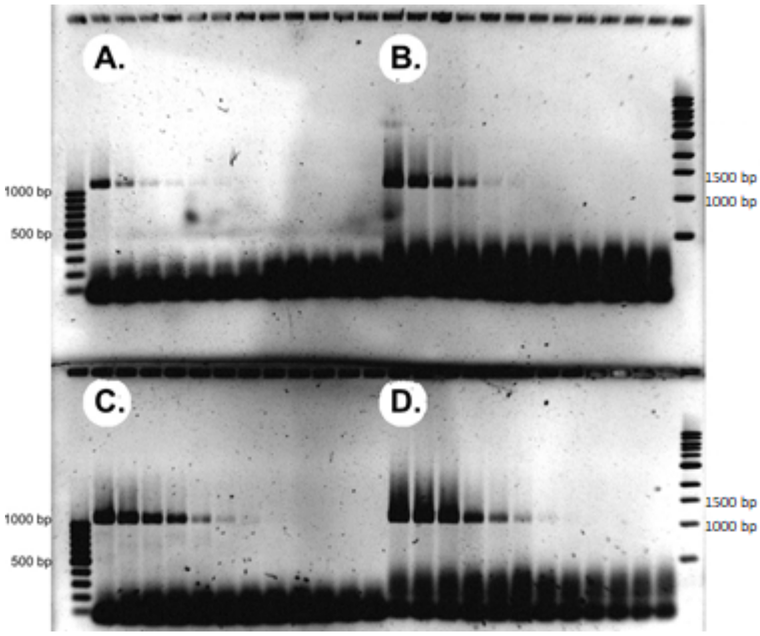

**Figure S2.** The influence of viral load on the amplification of 1,200 bp amplicon was analysed by agarose gel electrophoresis using samples, where RNA was serially diluted prior to cDNA synthesis and semiquantitative multiplex PCR (A, B) or quantitative multiplex PCR (C, D). Multiplex primer pool 1 (A, C) and pool 2 (B, D) were used in separate single tube reactions. (A-D.) From left to right: Decreasing viral loads (dilution factors $2^{0}$-$2^{-10}$ plus no template control). The uncropped image shows the results of agarose gel electrophoresis after 1,200 bp amplicon generating multiplex PCR. After image acquisition the colors were inverted for better visualization.

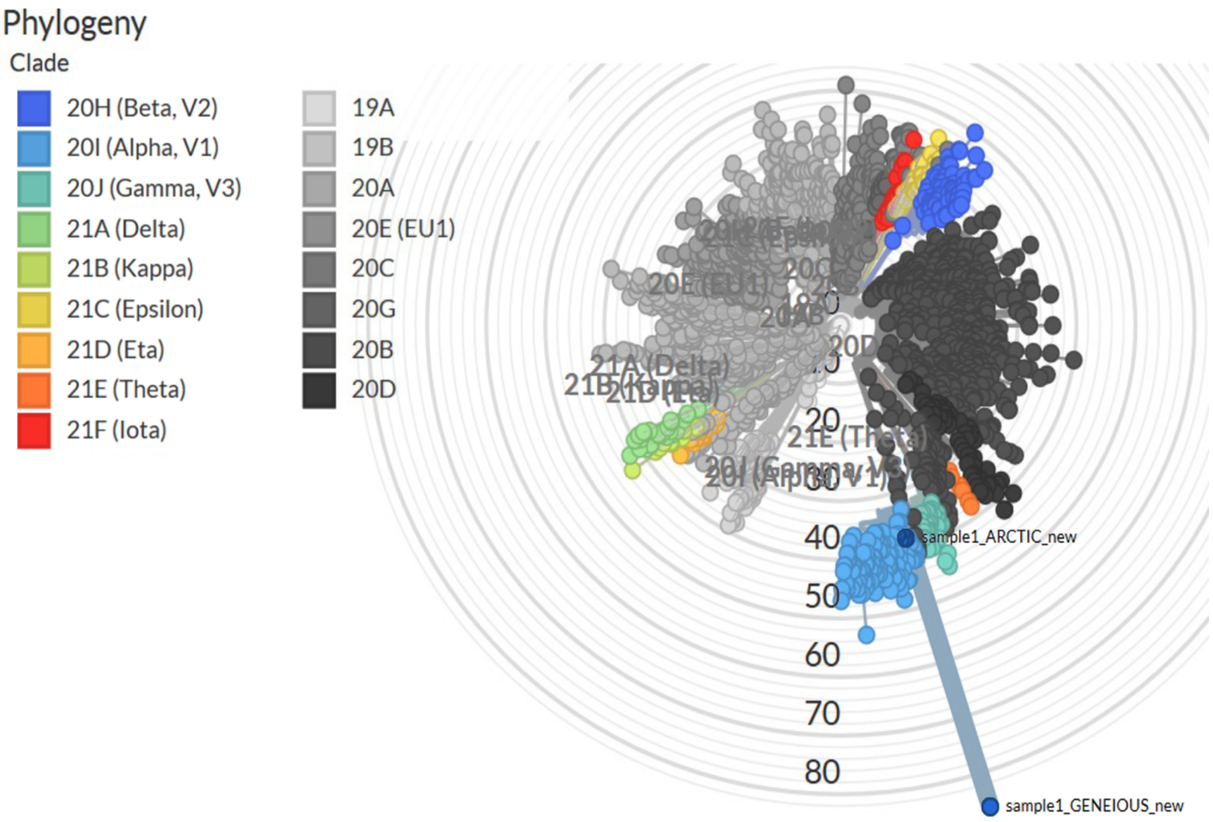


**Figure S3.** Comparison on the phylogenetic distance, when data from the same specimen was processed by 2 different bioinformatics pipelines Phylogenetic tree visualization was done using the Nextstrain web app (https://clades.nextstrain.org) for pathogen genome data analyses (Hadfield, Megill et al. 2018).

**Supplementary Information 1**

```
#!/usr/bin/env python3
# -*- coding: utf-8 -*-
"""
Created on Tue Jun 29 21:10:17 2021

@author: Erwan Sallard
"""
import sys
```

```
from Bio import Align
from Bio import SeqIO

medaka_consensus = sys.argv[1]
nanopolish_consensus = sys.argv[2]
output_folder = sys.argv[4]
coronavirus_reference_sequence = sys.argv[3]

output_suffix=medaka_consensus.split('/')[-1].split('.')[-3].split('_')[-1]

### Merging the consensus sequences

# when a sequence contains 'N's and the other solved the variant, we will take the solved
sequence.
# In the very rare cases where both algorithms disagree and none of them give 'N's, we give
priority to nanopolish

## perform the alignment
n_consensus = SeqIO.read(nanopolish_consensus,'fasta').seq
m_consensus = SeqIO.read(medaka_consensus,'fasta').seq
aligner = Align.PairwiseAligner()
aligner.mismatch_score = -1
aligner.target_open_gap_score = -1.53
aligner.target_extend_gap_score = -1
aligner.query_extend_gap_score = -1
aligner.query_open_gap_score = -1.53
alignment = str(next(aligner.align(n_consensus, m_consensus))).split('\n')

## replace 'N's in the nanopolish consensus by the medaka sequence if it is solved
'''
gérer blocs modifiés, ex 'NN-' vs '--T'
'''
aligned_nanopolish=alignment[0]
aligned_medaka=alignment[2]
merged_consensus=''
position=0
alignment_length=len(aligned_nanopolish)
while position<alignment_length:
    if aligned_nanopolish[position]=='N':
        # there is an unsolved sequence in the nanopolish alignments
        # first step: identify the extent of the unsolved region ('N's and '-'s)
        unsolved_begin = position
        unsolved_end = position
        while (unsolved_begin>1) and (aligned_nanopolish[unsolved_begin-1] in ['N','-']):
            unsolved_begin-=1
        while (unsolved_end<alignment_length-1) and (aligned_nanopolish[unsolved_end+1] in
['N','-']):
            unsolved_end+=1
        # second step: see if medaka solved the region
        medaka_segment=aligned_medaka[unsolved_begin:unsolved_end+1]
        medaka_solved=''
        for base in medaka_segment:
            if base in ['A','G','C','T']:
                medaka_solved+=base
        # third step: if it exists, append the solved sequence to the merged consensus
        if medaka_solved!='':
            merged_consensus+=medaka_solved
        else:
            # medaka too didn't solve the sequence, so we keep nanopolish's sequence
            merged_consensus+=aligned_nanopolish[unsolved_begin:unsolved_end+1]
        position=unsolved_end+1
    else:
        merged_consensus+=aligned_nanopolish[position]
        position+=1

## remove '-'
contiguous_sequences=merged_consensus.split('-')
merged_consensus=''
for segment in contiguous_sequences:
    merged_consensus+=segment

## save the consensus sequence in fasta format
fasta = open(output_folder+'/merged_consensus_'+output_suffix+'.fa','w')
fasta.writelines('>Merged_consensus_from_nanopolish_and_medaka_'+output_suffix+'\n')
consensus_length = len(merged_consensus)
line_number = int(consensus_length/60)+1
for line in range(line_number-1):
```

```
    fasta.writelines(merged_consensus[60*line:60*(line+1)]+'\n')
fasta.writelines(merged_consensus[60*(line_number)-1:])
fasta.close()


### Identifying all solved variants in the combined algorithms output

## align the merged consensus with the reference SARS-CoV-2
reference = SeqIO.read(coronavirus_reference_sequence,'fasta').seq
consensus = SeqIO.read(output_folder+'/merged_consensus_'+output_suffix+'.fa','fasta').seq
alignment = str(next(aligner.align(consensus, reference))).split('\n')
aligned_consensus=alignment[0]
aligned_reference=alignment[2]

## identify the solved variants in the consensus
variants_summary = []
# this list will record for all variants their position in the reference genome,
# and the reference and variant alleles
index, position = 0, 1
reference_length = len(reference)
alignment_length = len(aligned_reference)
while index < alignment_length:
    if aligned_reference[index]=='-' and aligned_consensus[index-1]!='N':
        # the consensus contains an insertion (and not an unsolved region)
        mismatch_end = index+1
        while mismatch_end<alignment_length and aligned_reference[mismatch_end]=='-':
            mismatch_end+=1
        variants_summary.append([position-1,aligned_reference[index-
1],aligned_consensus[index-1:mismatch_end]])
        index=mismatch_end
    elif aligned_consensus[index]=='-' and aligned_consensus[index-1]!='N':
        # the consensus contains an deletion (and not an unsolved region)
        mismatch_end = index+1
        while mismatch_end<alignment_length and aligned_consensus[mismatch_end]=='-':
            mismatch_end+=1
        variants_summary.append([position-1,aligned_reference[index-
1:mismatch_end],aligned_consensus[index-1]])
        position+=mismatch_end-index
        index=mismatch_end
    elif aligned_consensus[index]!=aligned_reference[index] and aligned_consensus[index]!='N'
and aligned_consensus[index-1]!='N':
        # the consensus contains a substitution (and not an unsolved region)
        mismatch_end = index+1
        while mismatch_end<alignment_length and
aligned_consensus[mismatch_end]!=aligned_reference[mismatch_end]:
            mismatch_end+=1
        reference_allele = aligned_reference[index:mismatch_end]
        while (reference_allele[-1]=='-'):
            # in case the substitution is followed by an insertion, there is no need to record
the '-' sign
            reference_allele = reference_allele[:-1]
            position-=1
        consensus_allele = aligned_consensus[index:mismatch_end]
        while (consensus_allele[-1]=='-'):
            consensus_allele = consensus_allele[:-1]
        variants_summary.append([position,reference_allele,consensus_allele])
        position+=mismatch_end-index
        index=mismatch_end
    else:
        index+=1
        position+=1

## create a .vcf file to save the variants and write the header
output_vcf = output_folder+'/merged_variants_'+output_suffix+'.vcf'
variants=open(output_vcf,'w')
variants.writelines('##fileformat=VCFv4.2\n')
variants.writelines('#CHROM\tPOS\tID\tREF\tALT\tQUAL\tFILTER\tINFO\n')

## fill the .vcf file
for variant in variants_summary:
    variants.writelines('SARS-CoV-
2_genome\t'+str(variant[0])+'\t.\t'+variant[1]+'\t'+variant[2]+'\t.\t.\t.\n')

variants.close()
```
